# Genetic and epigenetic architecture of mass spectrometry-derived lipids in blood and their role in lifetime Major Depressive Disorder

**DOI:** 10.64898/2026.09.02.26362049

**Authors:** Hannah M. Smith, Anne Richmond, Mark J. Adams, Asger Wretlind, Riccardo E. Marioni, Cristina Legido-Quigley, Andrew M. McIntosh

## Abstract

Major depressive disorder (MDD) is a highly prevalent psychiatric disorder that remains challenging to treat effectively. Difficulty predicting who will develop the disorder and which individuals will respond to treatment is most likely due to our limited mechanistic understanding and the lack of reliable biomarkers. The current literature suggests that lipid profiles differ in MDD; however, most studies do not integrate other omics layers to elucidate their potential causal relationship at the molecular level. Using phenotypic, genetic, epigenetic and mass-spectrometry lipid data from a Scottish-based cohort of approximately 1,000 participants, we examined associations with individual lipid species. We identified 77 common genetic variant associations (β range = -0.46 to 0.69, p < 3.1x10^-10^) and 82 genome-wide DNA methylation associations (β range = -1.88 to 1.39, p < 2.2x10^-10^). We identified 23 lipid species associated with lifetime MDD (β range = - 0.23 to 0.2, p < 0.05), including lipid classes such as lysophosphatidylcholine (LPC), diacylglycerol (DG) and phosphatidylcholine (PC). Our integration analysis showed that genetic variants and DNA methylation at sites annotated to the FADS gene cluster were linked with lipid species that were also associated with lifetime MDD status, including LPC(14:1) (β = 0.15, p = 0.03) and PC(19:0)/(20:4) (β = -0.14, p < 0.05). Our findings provide further support for the role of the FADS gene cluster in lipid metabolism and its association with MDD, while introducing DNA methylation as an additional molecular layer supporting this link.

## Introduction

Major depressive disorder (MDD) is a complex psychiatric disorder characterised by low mood and loss of interest ^1^. It affects approximately 332 million individuals globally and is a leading cause of disability ^1,2^. Despite its prevalence and impact, the underlying biology of MDD remains poorly understood, limiting effective risk prediction, prevention and treatment. A deeper understanding of the molecular changes that occur in MDD may help to identify those at increased risk early or provide potential novel therapeutic targets.

Emerging evidence suggests that lipid profiles in blood may play an important role in MDD. Individuals with MDD exhibit altered circulating lipid profiles, most consistently elevated triglycerides, whilst findings for very-low-density lipoprotein (VLDL) and high-density lipoprotein (HDL) levels have been less consistent across studies ^3,4^. Mass spectrometry-based lipidomics has also revealed associations between MDD and specific lipid classes, including sphingomyelins, glycerophospholipids and lysophospholipids ^5–7^. Genetic analyses using Mendelian randomisation and colocalization have also reported potentially causal associations between fatty acids and MDD risk in the UK Biobank ^8^. Together, these findings indicate that lipid dysregulation may be causally relevant to MDD.

Genome-wide association studies (GWAS) and epigenome-wide association studies (EWAS) provide complementary approaches for interrogating the molecular architecture of complex traits. GWAS can reveal genetic variants that influence lipid metabolism ^9^. EWAS can capture DNA methylation (DNAm) at cytosine-phosphate-guanine (CpG) sites in the genome that may reflect regulatory mechanisms or environmentally mediated biological responses ^10,11^. Common cholesterol measures, such as HDL and LDL are only weakly linked at the genetic level to many specific lipid molecules, highlighting the value of studying lipids in more detail ^12^. Studies have reported associations between DNAm at CpG sites with traditional clinical lipids and nuclear magnetic resonance spectroscopy (NMR) derived lipid measures ^13,14^. To our knowledge, no EWAS of mass spectrometry-derived lipid species have been published previously.

Most existing studies investigating the relationship between genetics, epigenetics, and MDD with lipids have done so in isolation. This fragmented approach limits our ability to identify shared biological pathways, making it more challenging to understand how lipid metabolism links to depression risk. Integrating genomic and epigenomic analyses with high-resolution lipidomic data offers an opportunity to bridge this gap and identify molecular architecture linking lipids to MDD.

In this study, we first conducted GWAS and EWAS of 565 blood-based lipid species in the Stratifying Resilience and Depression Longitudinally (STRADL) subset of the Generation Scotland cohort (sample size range = 664 to 1,058) ^15^. We then tested associations between individual lipid species and lifetime MDD (median n_cases_ = 255, median n_controls_ = 596) and integrated these results with the genomic and epigenomic findings. Our study aims to provide a more comprehensive understanding of the molecular architecture associated with lipid species that were linked to lifetime MDD.

## Methods

### Generation Scotland and STratifying Resilience and Depression Longitudinally

Generation Scotland is a family-based study of 24,084 individuals living in Scotland at the time of recruitment, which took place between 2006 and 2011 ^16,17^. Clinical, sociodemographic, genetic, epigenetic and proteomic data are available for participants of the study. In this study, we used data from a depression-focused investigation of a subset of Generation Scotland known as the STRADL ^15^ cohort, who were recruited for in-person testing between 2015 and 2019. The STRADL cohort is 59% female and has a median age of 62 years.

### Lipidomics

Lipid species were measured using liquid chromatography-mass spectrometry (LC-MS) and previously established methods^18,19^. Lipids were extracted with a two-phase extraction using methyl tert-butyl ether (MTBE) and water, and samples were analysed on a XEVO QTOF (Waters, US) in both positive and negative ionisation mode. Mass and retention time were compared to a library of expected measurements and pure internal standards representing each lipid family to identify molecules. Individual visual inspection of each chromatogram was used to flag or remove low-quality measures. Additional quality control was performed by checking for missingness and outliers in R v. 4.5.3. Lipids with more than 20% missing values were excluded, and extreme values were winsorized to the 0.01st–99.99th percentile (∼ 4SD). Normalisation was carried out using an isotope-labelled exogenous internal standard introduced into each blood sample. A batch correction was performed using the sample tray number to account for between-batch drift that can occur on longer MS runs. Positive and negative ionisation datasets were merged for downstream analysis.

### Genotyping

The Illumina HumanOmniExpressExome-8v1-2_A or HumanOmniExpressExome-8v1_A arrays were used for genotyping of Generation Scotland participants. Steps that were taken to ensure the genotype data were of high quality have been described previously ^20^. In brief, SNPs with high missingness (< 2% across all samples), a minor allele frequency (MAF) of < 1% and a Hardy-Weinberg p-value of < 1x10^-6^ were removed. Samples with high missingness were removed using a filter of < 2% across SNPs. In instances where genetically determined sex mismatched with reported sex, samples were excluded from analysis. Genetic outliers were identified as those more than six SD from the mean of the first two principal components upon integration with the 1,000 Genomes population ^21^. These quality control steps resulted in a dataset with 604,858 SNPs and 20,032 participants ^22^. Imputation was carried out on the SNP data using the Haplotype Reference Consortium (HRC) imputation panel ^23^. Imputation resulted in 24,161,581 genetic variants with MAF > 1% and INFO score > 0.4.

### DNA methylation

The measurement of DNA methylation in STRADL and the wider Generation Scotland cohort has been described in detail previously ^24^. Genome-wide DNA methylation (DNAm) was measured in STRADL using the Illumina Human-MethylationEPIC v1 BeadChip array. Processing of the data was carried out in two sets (N_set1_ = 504, N_set2_ = 372) using R ^25^. Samples were removed if DNAm-predicted sex did not match reported sex using Meffil ^26^. Samples were also removed if >0.5% of CpG sites had a detection p-value > 0.01, median signal intensity > 3 SD lower than expected and/or there was evidence of dye bias ^26^. Outliers were removed based on visual inspection of the log median intensity of methylated versus unmethylated signals per array generated using shinyMethyl ^27^. Probes were removed using Meffil if they had a beadcount < 3 in > 5% of samples and/or if they had a detection p-value > 0.01 in > 1% of samples ^26^. Outliers were also removed by visual inspection of multidimensional scaling (MDS) plots. According to X chromosome DNAm levels, 40 males were identified as outliers and removed from the dataset. Following re-normalisation of the data, it was confirmed that no outliers remained via MDS plots. The dasen method from the watermelon package ^28^ was used for normalisation, and the beta2m function in lumi ^29^ was used to convert to M values. Mean imputation was used to handle three missing methylation values in the dataset. There were 774,073 probes remaining for 876 individuals after quality control.

### Major depressive disorder

An individual was classified as having MDD if they met a Diagnostic and Statistical Manual of Mental Disorders (DSM) or an electronic health record code of depression-based diagnosis. DSM-based diagnoses were based on the Structured Clinical Interview for DSM disorders (SCID) at the baseline Generation Scotland appointment or from a composite international diagnostic interview (CIDI) at STRADL follow-up. For electronic health record diagnosis, hospital inpatient (SMR01 from 1980 to 2022), mental inpatient (SMR04 from 1980 to 2020) and general practitioner (GP, from 1980 to 2020) records were examined. GP records are only available for approximately 40% of Generation Scotland participants due to consent restrictions with individual GP surgeries. Individuals who had a diagnosis or record of schizophrenia or bipolar disorder were removed from both the cases and the control group before analysis.

### Genome-wide association analysis

Individual GWAS for each rank-based inverse normalised lipid species were performed using fastGWA, a linear mixed effects model method in GCTA ^30^. This method accounts for family structure by fitting a sparse genetic relationship matrix (GRM) as a random effect, generated using a cut-off value of 0.05. Age, sex and 10 genetic principal components (to capture population structure) were included as fixed effects covariates. Genetic variants with a minor allele frequency < 5% were excluded from analysis. Genomic inflation factors were calculated for each lipid species using the median χ² statistic divided by the expected median of the chi-square distribution under the null hypothesis. Genome-wide significance was defined as p < 5x10^-8^. Joint and conditional analysis was performed using the GWAS summary statistics for each lipid species using GCTA-COJO ^30,31^ to identify lead independent associations. Default settings were used for COJO, including a stepwise selection procedure across 10Mb regions and SNP linkage disequilibrium set to a cut-off of R^2^ < 0.9, avoiding multicollinearity. The COJO findings were considered genome-wide significant if p < 5x10^-8^. We also applied a more stringent Bonferroni-adjusted threshold of p < 3.1x10^-10^ to account for multiple testing. This threshold was determined by dividing the genome-wide significance threshold of 5x10^-8^ by 161, which was the number of principal components taken to explain 80% of the variance in the lipidomics data. HRC imputed data from Generation Scotland was used as the LD reference panel. Significant COJO SNPs were annotated to the nearest protein-coding gene using biomaRt (Ensembl GRCh37) ^32^. For each variant, the distance to each gene was calculated based on the genomic coordinates, with variants falling within a gene assigned a distance of 0 base pairs. The nearest protein-coding gene within ±50 kb of each variant was reported; variants without a protein-coding gene within this window were recorded as having no nearby gene. The functional consequences of variants identified as significant in the GWAS and COJO analyses were annotated using Ensembl Variant Effect Predictor (VEP; version 115) against the GRCh37 human reference genome ^33^. The most severe predicted consequence for each variant was retained for downstream analyses. Demographics for the participants included in the GWAS can be observed in **Table S1**.

### Epigenome-wide association analysis

EWAS analysis was carried out using linear regression with the fast linear function in the omics-data-based complex trait analysis (OSCA) software package ^34^. The methylation dataset was pre-regressed for age, sex, DNAm-estimated white blood cell proportions (WBCs), batch, set, an epigenetic smoking score and body mass index (BMI, measured in kg/m^2^) using linear regression. The residuals from each model were scaled (mean = 0, standard deviation = 1) before being used as predictors in the EWAS. WBCs were estimated using the Houseman method ^35^. The epigenetic smoking score was calculated using the smoking score (SSc) method from the EpiSmokEr R package ^36^. Lipids were pre-regressed for age, sex and relatedness using linear mixed effects models (lmekin function from coxme R package), and the rank-based inverse normalised residuals for each lipid were used as outcomes in the EWAS. A kinship matrix was constructed using the R package kinship2 ^37^ and included in the linear mixed effects models as a random effect. An epigenome-wide significance threshold was defined as < 3.6x10^-8^, as suggested by Saffari *et al* ^38^. Similar to the GWAS analyses, we accounted for multiple test correction by dividing the epigenome-wide significance threshold by the number of PCs taken to explain 80% of the variance in the lipidomics (3.6x10^-8^ / 161 = 2.6x10^-10^). CpG sites were annotated to genes using the IlluminaHumanMethylationEPICanno.ilm10b4.hg19 Bioconductor package ^39^. Demographics for the participants included in the EWAS can be observed in **Table S2**.

### Lipid associations with major depressive disorder

Linear mixed effects models using the lmekin function from the Coxme ^40^ R package were used to test associations between MDD status (predictor) and lipid species (outcome). Models were initially adjusted for age, sex and relatedness (median n_cases_ = 289, median n_controls_ = 681), followed by an additional adjustment for smoking status and BMI (median n_cases_ = 255, median n_controls_ = 596). Relatedness was modelled in the same way as in the EWAS analyses using a kinship matrix. Associations were considered nominally significant if p < 0.05. As with the GWAS and EWAS analyses, we also applied a more stringent Bonferroni-adjusted threshold of p < 0.0003 (0.05 / 161) to account for multiple testing. Demographics for the participants included in the MDD analysis can be observed in **Table S3**.

### Sensitivity analysis for lipid-regulating drugs

A sensitivity analysis for the GWAS, EWAS and MDD models was performed, additionally adjusting for whether an individual was taking a lipid-regulating drug within three months before blood draw. A list of lipid-regulating drugs was derived using British National Formulary (BNF) code 2.12, and participants’ electronic prescribing records were checked to determine if they were taking any medication within this category. A binary medication (yes/no) variable was generated using this information and included as a covariate in all sensitivity models. Of the 1,117 participants with lipids and genetic data available, 209 individuals were taking a lipid-regulating drug within the three months preceding their blood draw appointment.

## Results

### Genome-wide association study of lipids

GWAS analyses of 565 blood-based lipid species were performed using data from a median of 1,055 (range = 850 to 1,058) individuals in the STRADL subset of Generation Scotland, accounting for relatedness (via genetic relationship matrix), age, sex and 10 genetic PCs. Common genetic variants (MAF ≥ 5%) were tested for association with each lipid species. We observed 5,668 genome-wide significant (p < 5x10^-8^) SNP associations with 137 lipids (**Table S4**). A median genomic inflation factor of 1.01 (interquartile range = 1.003 to 1.05) was observed across lipid species (**Table S5**).

Next, we performed a conditional and joint analysis to identify independent loci associated with each lipid. SNPs with the lowest p-value within a locus were defined as the lead variant. We observed 77 independent associations with 71 lipid species (β range = -0.46 to 0.69, p < 5x10^-8^, **Table S6**). 92% of lipids had a single independent signal, and only a small number had two signals. SNPs were annotated to the nearest protein-coding gene (±50kb). The gene with the most lipid associations was *FADS2* with 28 lipids, followed by *ALDH1A2* and *SPTLC3,* each with 12 lipids (**Table 1**). Application of a Bonferroni adjusted threshold of p < 3.1x10^-10^, did not alter the significant findings from the conditional and joint analysis. Functional annotation of the 24 unique lead variants from the 77 independent associations was performed using the Ensemble Variant Effect Predictor (VEP), which showed that most variants were located in non-coding regions. The majority of variants were annotated as intronic (17/24), with additional variants located upstream (3/24), intergenic (2/24) and downstream (1/24). One variant was annotated as a synonymous variant. No missense or nonsense variants were identified among the lead variants.

**Table 1.**
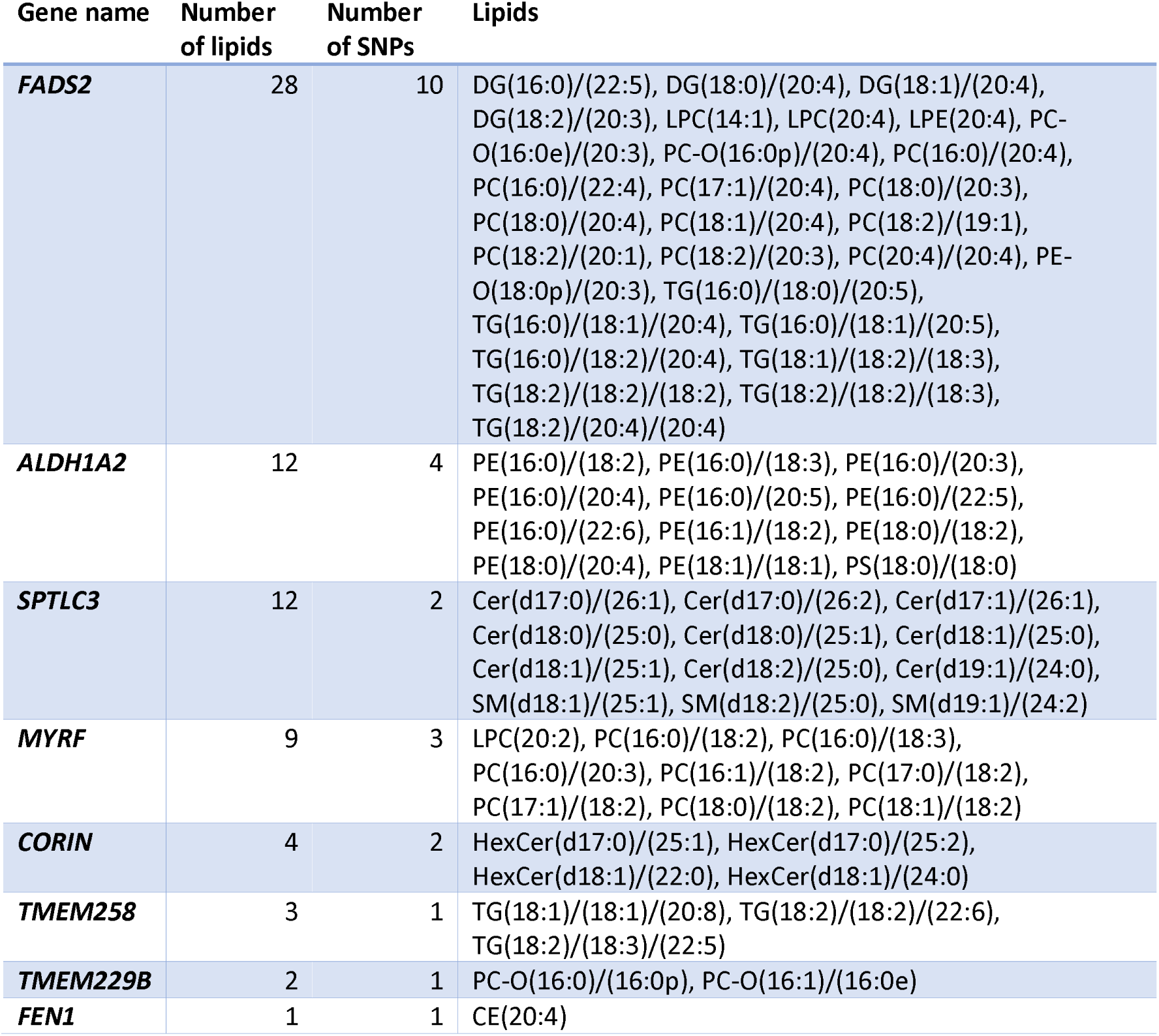
Summary of genetic variant associations with blood-based lipids from the conditional and joint analysis.

| Gene name | Number of lipids | Number of SNPs | Lipids |
| --- | --- | --- | --- |
| **FADS2** | 28 | 10 | DG(16:0)/(22:5), DG(18:0)/(20:4), DG(18:1)/(20:4), DG(18:2)/(20:3), LPC(14:1), LPC(20:4), LPE(20:4), PC-O(16:0e)/(20:3), PC-O(16:0p)/(20:4), PC(16:0)/(20:4), PC(16:0)/(22:4), PC(17:1)/(20:4), PC(18:0)/(20:3), PC(18:0)/(20:4), PC(18:1)/(20:4), PC(18:2)/(19:1), PC(18:2)/(20:1), PC(18:2)/(20:3), PC(20:4)/(20:4), PE-O(18:0p)/(20:3), TG(16:0)/(18:0)/(20:5), TG(16:0)/(18:1)/(20:4), TG(16:0)/(18:1)/(20:5), TG(16:0)/(18:2)/(20:4), TG(18:1)/(18:2)/(18:3), TG(18:2)/(18:2)/(18:2), TG(18:2)/(18:2)/(18:3), TG(18:2)/(20:4)/(20:4) |
| **ALDH1A2** | 12 | 4 | PE(16:0)/(18:2), PE(16:0)/(18:3), PE(16:0)/(20:3), PE(16:0)/(20:4), PE(16:0)/(20:5), PE(16:0)/(22:5), PE(16:0)/(22:6), PE(16:1)/(18:2), PE(18:0)/(18:2), PE(18:0)/(20:4), PE(18:1)/(18:1), PS(18:0)/(18:0) |
| **SPTLC3** | 12 | 2 | Cer(d17:0)/(26:1), Cer(d17:0)/(26:2), Cer(d17:1)/(26:1), Cer(d18:0)/(25:0), Cer(d18:0)/(25:1), Cer(d18:1)/(25:0), Cer(d18:1)/(25:1), Cer(d18:2)/(25:0), Cer(d19:1)/(24:0), SM(d18:1)/(25:1), SM(d18:2)/(25:0), SM(d19:1)/(24:2) |
| **MYRF** | 9 | 3 | LPC(20:2), PC(16:0)/(18:2), PC(16:0)/(18:3), PC(16:0)/(20:3), PC(16:1)/(18:2), PC(17:0)/(18:2), PC(17:1)/(18:2), PC(18:0)/(18:2), PC(18:1)/(18:2) |
| **CORIN** | 4 | 2 | HexCer(d17:0)/(25:1), HexCer(d17:0)/(25:2), HexCer(d18:1)/(22:0), HexCer(d18:1)/(24:0) |
| **TMEM258** | 3 | 1 | TG(18:1)/(18:1)/(20:8), TG(18:2)/(18:2)/(22:6), TG(18:2)/(18:3)/(22:5) |
| **TMEM229B** | 2 | 1 | PC-O(16:0)/(16:0p), PC-O(16:1)/(16:0e) |
| **FEN1** | 1 | 1 | CE(20:4) |

A sensitivity analysis was performed adjusting for whether participants were taking lipid-regulating medication within three months before blood draw. We observed 180 associations with 151 lipids in the conditional and joint analysis after adjusting for medication status (β range = -0.53 to 0.69, p < 5x10^-8^, **Table S7**). Of those 180 associations, 67 were also observed in the main GWAS at p < 5x10^-8^, all with the same direction of effect.

### Epigenome-wide association study of lipids

EWAS analysis of the 565 lipids was performed using data from a median of 806 participants in STRADL (range = 664 to 808), adjusting for age, sex, relatedness, WBCs, BMI and an epigenetic smoking score. We observed 322 significant associations with 138 lipids at p < 3.6 x10^-8^ (**Table S8**). These results included 67 unique probes that mapped to 40 unique genes. The number of significant CpG associations was attenuated to 82 with 36 lipids after a Bonferroni threshold of p < 2.2 x10^-10^ was applied (β range = -1.88 to 1.39). These 82 associations were composed of 23 unique CpG sites annotated to 8 unique genes. A single CpG site (cg06500161) annotated to *ABCG1* associated with 15 lipid species, and a single CpG site (cg06690548) annotated to *SLC7A11* associated with 10 lipid species (**Table 2**). A median genomic inflation factor of 1.03 (interquartile range = 0.98 to 1.15) was observed for the EWAS results (**Table S9**). We additionally adjusted the EWAS models for lipid-regulating medication use within the three months preceding blood draw. Of the 278 associations observed in the sensitivity EWAS at p < 3.6 x10^-8^, 221 were also observed in the main EWAS at the same significance threshold, and all showed the same direction of effect (**Table S10**).

**Table 2.** Summary of CpG associations with blood-based lipids from the EWAS analyses.

| Gene | Number of lipids | Number of CpGs probes | Lipids |
| --- | --- | --- | --- |
| ***ABCG1*** | 11 | 1 | HexCer(d17:0)/(25:2), HexCer(d17:1)/(25:1), HexCer(d18:1)/(24:1), PC-O(16:0p)/(18:2), PC-O(16:1)/(16:0p), PG(18:0)/(20:2), SM(d18:1)/(26:2), SM(d18:2)/(14:0), SM(d18:2)/(24:0), SM(d18:2)/(24:2), SM(d18:2)/(26:1) |
| ***FADS2*** | 10 | 15 | DG(16:0)/(22:5), DG(18:1)/(20:4), DG(18:2)/(20:3), PC-O(16:0e)/(20:3), PC(16:0)/(22:4), PC(17:1)/(20:4), PC(18:0)/(20:4), PC(18:1)/(20:4), PC(18:2)/(20:3), PE-O(18:0p)/(20:3) |
| ***SLC7A11*** | 10 | 1 | CE(17:0), DG(18:0)/(18:0), PE(16:0)/(20:4), PS(18:0)/(18:0), TG(16:0)/(18:1)/(22:6), TG(16:0)/(18:2)/(22:5), TG(16:1)/(18:1)/(22:5), TG(18:1)/(18:1)/(20:5), TG(18:1)/(18:2)/(20:4), TG(18:2)/(18:2)/(20:3) |
| ***JDP2*** | 3 | 1 | CE(17:0), DG(18:0)/(18:0), PC(20:4)/(22:6) |
| ***LOC100132354*** | 2 | 1 | CE(17:0), DG(18:0)/(18:0) |
| ***TXNIP*** | 1 | 1 | CE(17:0) |
| ***FADS1*** | 1 | 1 | PC-O(16:0e)/(20:3) |
| ***MIR1908;FADS1*** | 1 | 1 | PE-O(18:0p)/(20:3) |

### Lipid associations with lifetime Major Depressive Disorder

We tested whether lipid profiles differed by lifetime MDD status (median n_cases_ = 289, median n_controls_ = 681), in linear mixed effects models adjusted for age, sex and relatedness, identifying nominally significant associations with 23 lipids (β range = - 0.23 to 0.2, p < 0.05, **Figure 1, Table S11**). No associations were significant at a Bonferroni-adjusted threshold of p < 0.0003. After further adjustment for BMI and smoking status (median n_cases_ = 255, median n_controls_ = 596), only two nominally significant associations were observed. MDD was positively associated with PI(18:1)/(22:6) and negatively associated with FA(26:2) (β = 0.15 and -0.16, respectively, p < 0.05, **Table S12**). After additional adjustment for lipid-regulation drug status, five lipids were nominally associated with lifetime MDD (β range = -0.16 to 0.17, p < 0.05, **Table S13**), with no associations observed at a Bonferroni-adjusted threshold of p < 0.0003.

**Figure 1:**
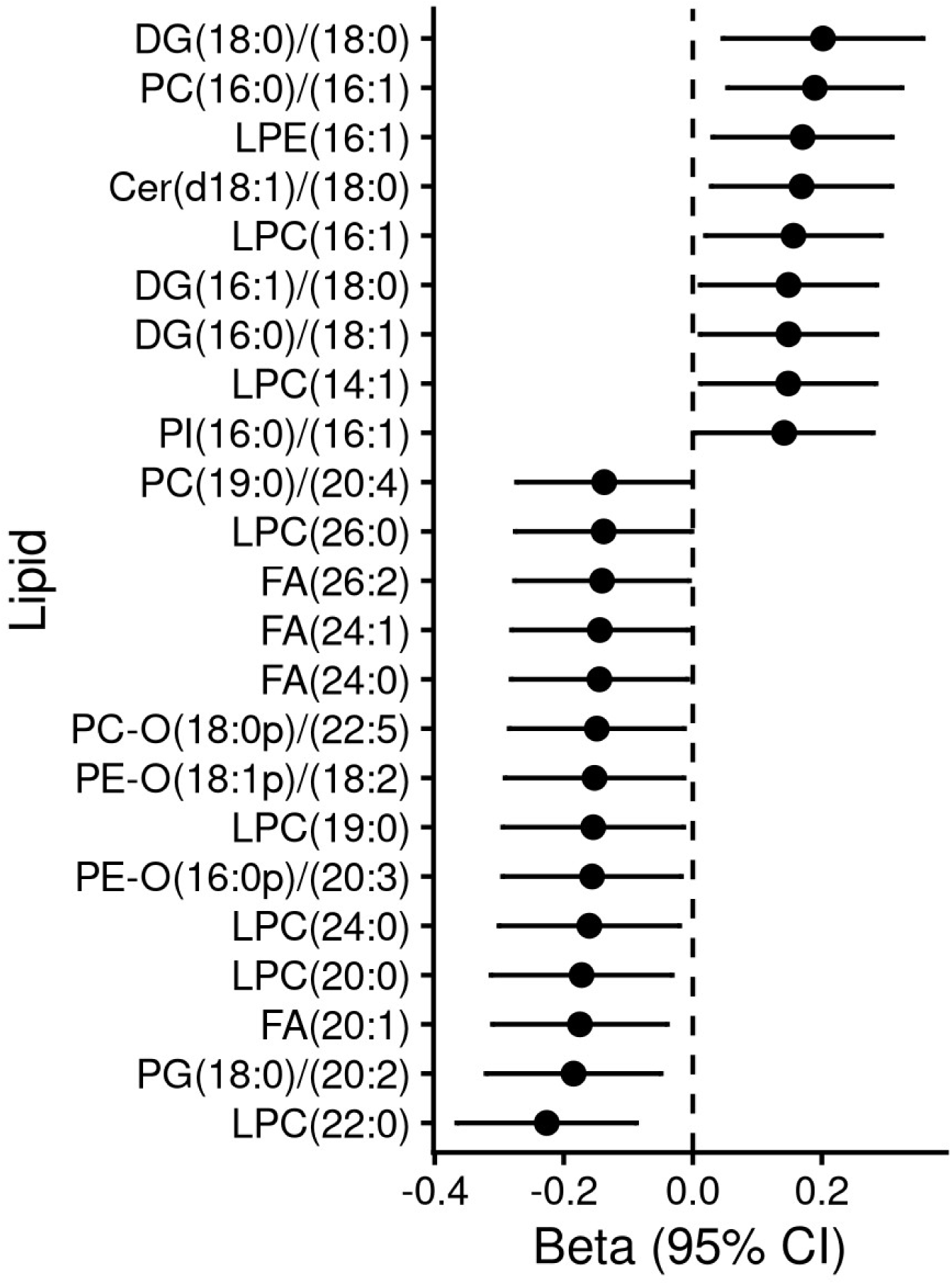
Lifetime MDD associations with blood-based lipids after accounting for age, sex and relatedness. Point estimates and 95% confidence intervals (95% CI) are shown for associations between life MDD and lipids for which p < 0.05.

### Integration of GWAS, EWAS and MDD associations

We subset our GWAS and EWAS significant findings to lipids identified as nominally significant (p < 0.05) in the MDD analysis adjusted for age, sex and relatedness (**Table 3**). One genetic variant association annotated to *FADS2* (11_61619829_C_A) was negatively associated with LPC(14:1) (β = -0.29, p = 2.2x10^-11^), a lipid species that was positively associated with MDD (β = 0.15, p = 0.03). DNAm levels at cg11250194 annotated to *FADS2* was positively associated with a different lipid species, PC(19:0)/(20:4) (β = 1.04, p = 3.3x10^-9^), that was found to negatively associate with MDD (β = -0.14, p < 0.05). Four lipids that were negatively associated with MDD: LPC(22:0), LPC(24:0), LPC(26:0) and PG(18:0)/(20:2) (βrange = -0.23 to -0.14, p < 0.05), were also found to be negatively associated with DNAm levels at cg06500161 annotated to *ABCG1* (β_range_ = -1.62 to -1.2, p < 2.2 x10^-10^).

**Table 3.** CpG associations with blood-based lipids that were associated with MDD at p < 0.05 in models adjusted for age, sex and family structure.

| Probe | Gene | beta | p | Lipid |
| --- | --- | --- | --- | --- |
| cg00574958 | *CPT1A* | -0.78876 | 2.99E-08 | DG(16:0)/(18:1) |
| cg06690548 | *SLC7A11* | -0.51263 | 1.52E-08 | DG(16:0)/(18:1) |
| cg00574958 | *CPT1A* | -0.78876 | 2.99E-08 | DG(16:1)/(18:0) |
| cg06690548 | *SLC7A11* | -0.51263 | 1.52E-08 | DG(16:1)/(18:0) |
| cg00574958 | *CPT1A* | -0.96928 | 4.64E-10 | DG(18:0)/(18:0) |
| cg26974062 | *TXNIP;NBPF20;NBPF10* | -0.77128 | 4.04E-09 | DG(18:0)/(18:0) |
| cg18120259 | *LOC100132354* | -1.48826 | 3.30E-11 | DG(18:0)/(18:0) |
| cg06088069 | *JDP2* | -1.85492 | 1.10E-11 | DG(18:0)/(18:0) |
| cg19693031 | *TXNIP* | -0.7061 | 1.53E-08 | DG(18:0)/(18:0) |
| cg06690548 | *SLC7A11* | -0.64192 | 1.21E-11 | DG(18:0)/(18:0) |
| cg05325763 | *CPT1A* | -0.72897 | 3.37E-08 | DG(18:0)/(18:0) |
| cg19884658 | *KLHL21* | -1.17673 | 1.09E-08 | DG(18:0)/(18:0) |
| cg26883048 | *MLLT10* | -0.6905 | 1.81E-08 | LPC(20:0) |
| cg06500161 | *ABCG1* | -1.19406 | 3.55E-08 | LPC(22:0) |
| cg06500161 | *ABCG1* | -1.35333 | 4.08E-10 | LPC(24:0) |
| cg06500161 | *ABCG1* | -1.2361 | 9.24E-09 | LPC(26:0) |
| cg06690548 | *SLC7A11* | -0.50467 | 1.10E-08 | PC(16:0)/(16:1) |
| cg11250194 | *FADS2* | 1.04214 | 3.31E-09 | PC(19:0)/(20:4) |
| cg06500161 | *ABCG1* | -1.6194 | 1.05E-14 | PG(18:0)/(20:2) |

## Discussion

In our study, we investigated genetic, epigenetic and lifetime MDD associations with circulating lipid species quantified using high-resolution mass spectrometry lipidomics in approximately 1,000 participants from the STRADL cohort.

Our GWAS analysis identified 77 common genetic variant associations with 71 lipids at p < 5x10^-8^, most with large effects (absolute median β = 0.36, β range = -0.46 to 0.69). Eight common genetic variants annotated to *FADS2* were associated with 28 lipid species. *FADS2* encodes an enzyme that is involved in the biosynthesis of long-chain polyunsaturated fatty acids from essential fatty acids ^41^. It is well established that genetic variants in the *FADS* gene cluster are known to influence the efficacy of dietary fatty acid metabolism, and thus several associations between variants in these genes and lipid species levels have been reported previously ^9,42–44^. We observed associations between a single genetic variant annotated to *FADS2* and LPC(14:1), a lipid species found to positively associate with MDD in our study. To our knowledge, an association with the short monounsaturated LPC(14:1) has not been reported previously, as associations are typically described between *FADS1*/*FADS2* and longer-chain fatty acids ^9^.

Epigenetic analyses further supported the involvement of variation within the molecular architecture of the *FADS* gene cluster in MDD-related lipid traits, with EWAS of lipids identifying a CpG site annotated to *FADS2* that was associated with lipid species linked to MDD. cg11250194, was positively associated with PC(19:0)/(20:4), a lipid species that was negatively associated with lifetime MDD, suggesting a potential epigenetic link between *FADS2*, lipid metabolism and depression. Our genetic (and epigenetic) findings converge on the same pathways as a recent large-scale metabolomics study in the UK Biobank that identified potentially causal links between polyunsaturated fatty acid levels and MDD risk, with genetic signals mapping to the *FADS* gene cluster ^8^.

Our EWAS analysis also revealed 11 lipid associations with a single methylation site (cg06500161) annotated to *ABCG1*, a gene that encodes a cholesterol transporter protein ^45^. Four of those lipids (LPC(22:0), LPC(24:0), LPC(26:0) and PG(18:0)/(20:2)) were also found to be associated with lifetime MDD in models adjusted for age, sex and relatedness. Methylation at this *ABCG1* CpG site is consistently associated with circulating cholesterol levels and BMI ^46–49^, which were observed after adjustment for BMI. Notably, none of the four lipid-MDD associations remained nominally significant after further adjustment for BMI and smoking, suggesting potential confounding or mediation by metabolic factors.

There was limited overlap between loci identified by the genetic and epigenetic approaches. The *FADS* cluster was the only region detected in both analyses. The epigenetic signals mapped to CpG sites at *ABCG1*, *TXNIP*, *SLC7A11* and *JDP2*, and, at the epigenome-wide threshold, *CPT1A*. Each has established associations with environmental and lifestyle exposures rather than with lipid levels; methylation at *ABCG1* is associated with adiposity and circulating cholesterol ^50^, *CPT1A* with fatty acid oxidation ^51^, *TXNIP* with glycaemia ^52^, and *SLC7A11* and *JDP2* with alcohol consumption ^53^. This pattern suggests partly separable influences on the circulating lipidome. A subset of the lipid species identified, including lipids with comparatively low abundance or less common acyl chain lengths, such as PC(19:0/20:4), may represent diet-derived or exogenously acquired fatty acids rather than products of endogenous metabolism. Similar species have been reported previously in comparable plasma lipidomic panels ^54^.

Key strengths of this study include the integration of GWAS and EWAS findings within the same cohort to explore potential molecular mechanisms linking blood-based lipid species to depression. The use of mass spectrometry-based lipidomics enabled detailed quantification of individual lipid species, providing substantially greater molecular resolution than standard metabolomic panels, which predominantly capture broader lipid measures ^55^. This high-resolution approach, to our knowledge, is the first epigenome-wide association study of circulating lipid species measured using mass spectrometry, offering new insights into the molecular architecture of lipid regulation. The relatively large sample size for a deeply phenotyped lipidomics dataset further enabled systematic investigation of lipid-genetic and lipid-epigenetic relationships and their potential relevance to lifetime MDD.

Whilst the current sample size precludes genetically informed causal inference methods due to an insufficient number of instruments per trait, this study suggests that further studies of mass spectrometry-derived lipids in MDD are likely to help elucidate the causal links between lipids and MDD. A further limitation is that the exclusive inclusion of individuals of European ancestry limits the generalisability of these findings. Future work should include larger, more ancestrally diverse cohorts to determine the replicability and translatability of these findings.

To conclude, this study provides further support for a relationship between polyunsaturated fatty acids and MDD, which is connected to variants in the *FADS* gene cluster. We add another molecular layer by showing that a lipid species, PC(19:0)/(20:4), is both associated with lifetime MDD and methylation levels at a CpG annotated to *FADS2*.

## Funding

H.M.S was funded by the Translational Neuroscience Transition Fund (**Wellcome**: 21843/Z/19/Z). H.M.S and A.M were funded by **AMBER (Wellcome** 226770/Z/22/Z). A.M was also funded by **WTIA (Wellcome** 220857/Z/20/Z)**, ImmunoMIND (UKRI** MR/Z50354X/1, MR/Z000548/1) and **Metabolic Psychiatry (**MR/Z503563/, MR/Z000548/). Generation Scotland was funded by a grant from the Chief Scientist Office of the Scottish Government Health Directorates (CZD/16/6) and the Scottish Funding Council (HR03006). Genotyping and DNA methylation profiling of the GS samples was carried out by the Genetics Core Laboratory at the Edinburgh Clinical Research Facility, University of Edinburgh, Scotland, and was funded by the Medical Research Council UK and the Wellcome Trust (Wellcome Trust Strategic Award ‘STratifying Resilience and Depression Longitudinally’ Reference 104036/Z/14/Z). DNA methylation profiling was also funded in part by the Wellcome Trust Investigator Award (Reference 220857/Z/20/Z). GS is currently supported by the Wellcome Trust (216767/Z/19/Z). M.J.A. is funded by **MRC LPS** (UKRI4951). Generation Scotland received core support from the **Chief Scientist Office** (CZD/16/6) and the **Scottish Funding Council** (HR03006) and funding from **Wellcome** (216767/ Z/19/Z). C.L.Q and A.W are funded by a Lundbeck Fonden grant (R344-2020-989).

## Ethics

This study involves human participants. All components of Generation Scotland received ethical approval from the NHS Tayside Committee on Medical Research Ethics (REC Reference Number: 05/S1401/89). Generation Scotland has also been granted Research Tissue Bank status by the East of Scotland Research Ethics Service (REC Reference Number: 25/ES/0013), providing ethical approval for a wide range of uses within medical research. All components of STRADL received formal, national ethical approval from the NHS Tayside committee on research ethics (reference 14/SS/0039). Written informed consent was obtained from all participants. This study was performed in accordance with the Helsinki declaration.

## Data access and availability

All code associated with this manuscript can be found in the following GitHub repository: https://github.com/HM-Smith/genetic_epigenetic_mdd_lipids. The full GWAS and EWAS summary statistics will be made available on publication. According to the terms of consent for Generation Scotland participants, access to data must be reviewed by the Generation Scotland Access Committee. Access requests can be made through the researcher portal (https://gsaccess.igc.ed.ac.uk) and normally take up to six weeks for approval. Further information can be found on the Generation Scotland website: https://genscot.ed.ac.uk/for-researchers/access.

## Conflicts of interest

CLQ has received consultancy fees from Pfizer. She has received honoraria, travel or speakers’ fees from Biogen and research funds from Pfizer, Novo Nordisk and Waters Corporation unrelated to this work. She is the director and founder of the consultancy company BrainLogia.

All other authors declare no conflict of interest.

